# Assessment of Environmental and Occupational Risk Factors for the Mitigation and Containment of a COVID-19 Outbreak in a Meat Processing Plant

**DOI:** 10.1101/2021.09.17.21262959

**Authors:** Nicola Walshe, Mehael Fennelly, Stig Hellebust, John Wenger, John Sodeau, Michael Prentice, Charlene Grice, Vincent Jordan, John Comerford, Vicky Downey, Carla Perrotta, Grace Mulcahy, Donal Sammin

## Abstract

Throughout the COVID-19 pandemic, meat processing plants have been vulnerable to outbreaks of SARS-CoV-2 infection. Transmission of the virus is difficult to control in these settings because of a combination of factors including environmental conditions and the specific nature of the work. This paper describes a retrospective outbreak investigation in a meat processing plant, a description of the measures taken to prevent or contain further outbreaks, and insights on how those with specific knowledge of the working environment of these plants can collaborate with public health authorities to ensure optimal outbreak control. The plant experienced 111 confirmed positive asymptomatic cases in total with an estimated attack rate of 38% during a five-week period. Four weeks after the first case, mass screening of all workers was conducted by the public health authorities. Thirty-two workers tested positive, of which 16 (50%) worked in one particular area of the plant, the boning hall (n=60). The research team prepared and carried out semi-structured interviews with the plant personnel who were charged with COVID control within the plant. They carried out assessments of operational risk factors and also undertook air quality monitoring in the boning hall and abattoir. The air quality measurements in the boning hall showed a gradual build-up of carbon dioxide and aerosol particles over the course of a work shift, confirming that this poorly ventilated area of the plant had an environment that was highly favourable for aerosol transmission of SARS-CoV-2. Assessment of operational conditions incorporated visual surveys of the plant during the working day. Prior to and during the first two weeks of the outbreak, multiple measures were introduced into the plant by management, including physical distancing, provision of educational material to workers, visitor restrictions, and environmental monitoring. After the implementation of these measures and their progressive refinement by plant management, the factory had no further linked cases (clusters) or outbreaks for the following 198 days. The tailored approach to risk mitigation adopted in this meat processing plant shows that generic risk mitigation measures, as recommended by public health authorities, can be successfully adapted and optimized by designated plant emergency response teams.

## 1 Introduction

Throughout the COVID-19 pandemic, meat processing plants (MPPs) have proved vulnerable to transmission of SARS-CoV-2, and outbreaks have affected their workers worldwide (1). Viral transmission is difficult to control in these settings because of a combination of factors including environmental conditions, the nature of the work, and difficulties in implementing physical distancing within meat plants (2). Clusters of COVID-19 have occurred in MPPs across the world - there are reports of such clusters appearing in the USA, Netherlands, Germany, France, the UK, and Australia^1^. Meat and poultry plants were heavily affected across the US, such that by the end of April 2020, the Centers for Disease Control and Prevention (CDC) had received reports of at least one COVID-19 plant outbreak in 19 of 23 states surveyed (3).

The potential risk factors in MPPs include high occupancy, the physically demanding nature of the work and environmental conditions which are unavoidably and deliberately – for food hygiene reasons – very different from those likely to be encountered in other work settings. Although the type of work and the working environment in MPPs may pose challenges in preventing the spread of an infectious disease like COVID-19 among workers, there is a large degree of variation in the extent to which plants have been able to reduce this risk. Workplace risk-mitigation measures have the potential to reduce the transmission risk and improve the containment of an outbreak once declared (4). Outbreaks in MPPs can be large and, if not limited, may result in virus spill-over back out into the community (2). There is mounting evidence that superspreading events play an important role in the establishment and maintenance of SARS-CoV-2 infection. A recent review, which simulated the potential of infected individuals to cause large numbers of secondary cases, highlighted how targeting locations where superspreading is most likely to happen, such as MPPs, could have a very significant impact in controlling virus spread (5). It is likely that MPPs will continue to be vulnerable to COVID-19, including superspreading events, in the face of potential future variants and the requirement for booster vaccination. Therefore, it is crucial to understand the specific features of these working environments that favour SARS-CoV-2 transmission, allowing for additional risk mitigation measures to be put place to help protect workers and the wider community from contracting COVID-19. Furthermore, this information will help improve preparedness for future emerging infectious respiratory diseases.

The first reported case of COVID-19 in Ireland was confirmed by a positive PCR test in late February 2020 (6). In the following weeks and months as further COVID-19 cases and deaths occurred, the government introduced a series of restrictions and regulations, eventually leading to a national lockdown. As farming and food production were identified as *essential services* under the COVID-19 regulations, MPPs in Ireland remained open throughout the pandemic, operating within guidelines provided by public health authorities. The agri-food industry accounts for ~8% of Ireland’s GDP and ~160,000 jobs^2^. The meat-processing industry is not only an important component of the agri-food sector, it also forms an important part of the national and international food supply. Therefore, not only from a public health point of view but also from an economic perspective, it is crucial to the industry, the workers, and the wider national community that investigation into potential risk factors for SARS-CoV-2 transmission, in Irish meat processing plants is conducted. This is important particularly in the context of new and potentially more transmissible variants of the virus.

Despite implementing guidelines provided by the public health authorities, numerous cases and outbreaks of SARS-CoV-2 infection have been detected amongst meat-processing staff in Ireland. This study describes a retrospective investigation of a COVID-19 outbreak in an Irish meat plant, that included semi-structured interviews with plant management, air quality assessments, an evaluation of operational factors and verification of risk mitigation measures.

## 2 Methods

### 2.1 Study Team

Arising from concerns on the vulnerability of MPPs in Ireland to COVID-19 outbreaks, the Department of Agriculture, Food and the Marine (DAFM), who are the oversight body for animal welfare and food safety in export meat plants, and have a permanent presence in these plants, agreed to co-ordinate a pilot study of operational and environmental factors influencing transmission of COVID-19 within a primary meat processing facility. A study team was assembled, comprising researchers from three universities, veterinary inspectors from DAFM, and an inspector from the Health and Safety Authority (HSA). The collective expertise of the study team included infection control, occupational health, aerosol science, meat plant operations and food hygiene, epidemiology, and public health. The team corresponded closely with public health doctors from the Health Services Executive (HSE), who were responsible for local outbreak control and provided anonymised PCR rest results, and key personnel from the local management team in the meat plant.

### 2.2 Study Design

DAFM performed an initial critique of the growing body of evidence from the emerging literature on SARS-CoV-2 and COVID-19. In addition, peer-reviewed reports on the transmission of severe acute respiratory syndrome (SARS) since 2002 and Middle East Respiratory Syndrome (MERS) since 2011. Recently published studies on the physics of transmission of respiratory infections were also considered.

The key considerations arising from this initial review of the literature informed both the study design and the composition of the investigative team. These included the increasing recognition of superspreading events giving rise to large numbers of cases of COVID-19 in various congregate settings and the likely importance of aerosol transmission in these events, as distinct from either droplet or contact transmission (1, 2, 7–13). The working assumption was that a superspreading event was likely to have occurred within meat plants in which large numbers of PCR positive workers were identified. This led to the hypothesis that conditions within meat plants may favour aerosol transmission of the virus and clarified the objective of the pilot study – to observe and measure operational and environmental factors that would support or refute this hypothesis by engaging in a retrospective investigation of a COVID-19 outbreak in a meat processing plant. The study design (Figure 1) included the collection of epidemiological, operational, and environmental data.

**Figure 1.**
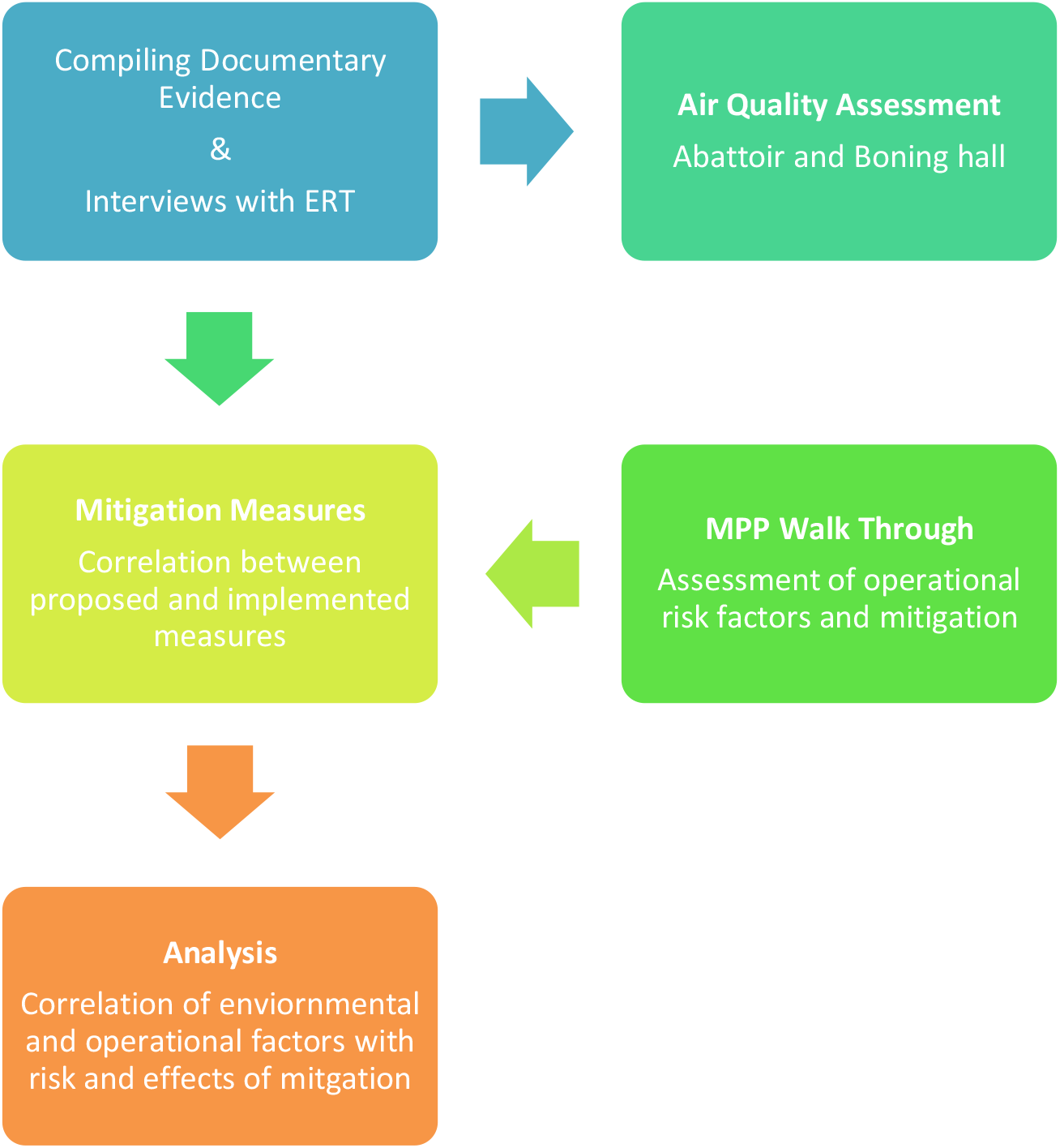
Study design. An interdisciplinary study team was assembled which undertook to compile relevant data and information from plant management, carry out environmental assessments, and observe operational factors and implementation of risk-mitigation measures directly. MPP – Meat processing plant. ERT - Emergency Response Team

The following criteria were used to select the plant: 1) That an outbreak of COVID-19 had occurred in the plant in early-mid 2020, 2) that mass PCR testing of the workforce had been undertaken as part of the outbreak investigation, 3) that a large proportion of the workforce had tested positive for the virus, and 4) that the plant comprised all aspects of primary processing of red meat at one site, including slaughter, meat-cutting or boning, packaging, and dispatch.

### 2.3 Data Collection

In advance of the study team visiting the site, all relevant documentation was assembled in consultation with plant management. A site visit for further in-depth investigation of operational risk factors, comprising: (a) a semi-structured interview with the plant Emergency Response Team (ERT) that had been established by plant management and (b) a walkthrough inspection of the facility to verify implementation of controls. On-site, continuous monitoring of air quality in selected areas of the plant was undertaken over a 10-day period.

The information gathered included details of the plant layout including spatial measurements pertaining to all aspects of production within the plant and environmental parameters related to ventilation and temperature control. Details of the workforce including total number employed and distribution throughout the factory departments were recorded, along with details of the shift assignment according to department. The sequence of COVID-19 related cases that were recorded by the factory were collated. The distribution of PCR positive cases throughout each area of the plant was mapped. Plant management provided documentation on the progressive implementation, and refinement of risk mitigation measures from the time at which public health concerns in relation to COVID-19 first began. This sequential implementation of risk assessment and risk mitigation measures before, during and after the outbreak was assessed.

The team also compiled a case log review gathering anonymised details for each worker who tested PCR positive for SARS-CoV-2; clinical presentation (i. e. if they presented with or without symptoms), location within the plant and details on possible links to or close contacts with other workers in the plant.

The interview was based on historical data, reflections of relevant management, the ERT, and current recommendations regarding risk on site. The period of interest extended from the date on which COVID-19 was recognised as a risk on site in early 2020 for approximately six months.

### 2.4 Air Quality Measurements

The areas for air quality assessment were pre-selected based on documented evidence on the proportion of the overall number of PCR-positive cases which had occurred in each area during the outbreak – notably a meat cutting room or “boning hall” with a relatively high proportion of the cases and an abattoir with a relatively low proportion of the cases. Indoor air quality measurements were conducted in these two areas of the plant over several days in August 2020. Carbon dioxide (CO_2_) concentrations, temperature and relative humidity were continuously recorded using an AirVisual Pro air quality monitor (IQAir, Staad, Switzerland). The number and size distribution of aerosol particles over the size range 0.75 – 12 μm was measured in real-time using a Wideband Integrated Bioaerosol Sensor (WIBS-4a; Droplet Measurement Technologies, Colorado, USA), which also uses fluorescence to identify the fraction of aerosol particles that are bioaerosols, i.e. of biological origin (13). Both instruments were enclosed in a cabinet with air sample intake at a height of 1.5 m above ground level and deployed in the boning hall for three days and in the abattoir for two days. The measured CO_2_ concentrations were averaged over 5 minute intervals, while the fluorescent and total particle counts were summed over the same 5 minute interval.

In a follow-up study, the performance of a newly-installed air filtration device (Camfil® CC6000 air cleaner; Camfil, Dublin, Ireland) was assessed in the same boning hall during the period 20-25 September 2020. The device was installed on a steel frame in the centre of the boning hall. The device is equipped with H14 HEPA filters, which are ≥ 99.9995% efficient at removing particles of 0.3 μm diameter. At full operating capacity, the device is able to filter 6000 m^3^ of air per hour. Continuous measurements of aerosol particles using the WIBS-4a were undertaken while the device was operating at either 50% or 100% capacity during the working shift and compared with the same measurements taken when the device was not operating.

### 2.5 Ethical Clearance

University College Dublin Human Research Ethics committee provided ethical clearance (**LS-E-20-196-Mulcahy)**. The name and location of the plant have been anonymized. All personal data was de-identified before being provided to the research team. All participants provided informed consent for inclusion in the study.

## 3. Results

### 3.1 Characteristics of the Outbreak

The MPP at the time of the outbreak had a total of 290 workers distributed across 20 different areas and departments. A worker in the boning hall was absent from work in early 2020. This person developed symptoms while absent from work and also tested positive six days later. They indicated that their spouse had also subsequently developed symptoms and tested positive for COVID-19. This was the first case in which it was suspected that infection had been acquired within the plant. It occurred approximately one month after the first case documented in Ireland, and two weeks after the first public health restrictive measures were introduced. By the end of March 2020, there were 2990 officially diagnosed cases of COVID-19 in Ireland, the number of admissions to hospital was 834 (27.9%) and the epidemic trajectory was increasing. The second symptomatic case was also a worker in the boning hall who presented with symptoms three days after the index case had been reported as symptomatic. An outbreak (defined by the HSE as two or more cases occurring within 14 days) was declared on this day. The third symptomatic case, also a worker in the boning hall, presented the following day. The fourth symptomatic case was from outside the boning hall, and had frequented a common area available to all MPP employees. Subsequent cases occurred in other areas of the plant, as outlined in Table 1. A summary timeline of detection of PCR-positive cases is shown in Figure 2.

**Table 1.** Number of workers testing PCR-Positive of a meat processing plant from initial mass testing until late 2020. The total number of workers in each production area is also given.

| Department | Number of PCR-Positive Cases/Total Workers |
| --- | --- |
| Boning Hall | 60 /111 |
| Dressing Line 1 | 15/30 |
| Dressing Line 2 | 7/42 |
| Other production areas | 17/29 |
| Other Non-production areas | 8/24 |

**Figure 2.**
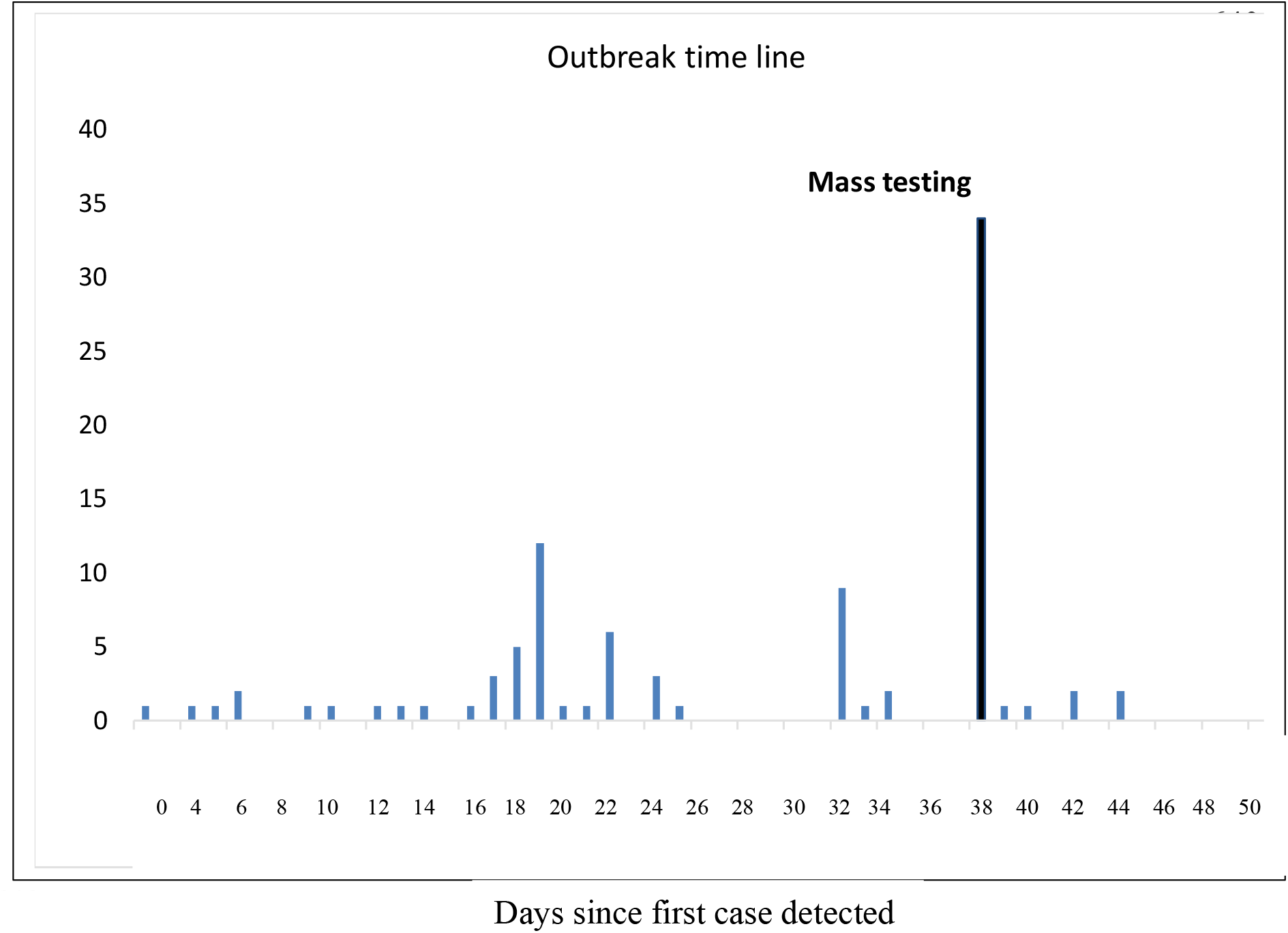
Time of detection and numbers of PCR-positive workers over the course of a COVID-19 outbreak in a meat processing plant during 2020. Forty-two PCR-positive cases shown from day 38 arose from mass PCR-testing of all workers on site.

Up to this time, national health policy recommendations stated that PCR testing was only needed for close contacts that were displaying symptoms of the disease (cough, shortness of breath or elevated temperature). However, as part of developing national policy, the MPP was advised by the local public health representatives that Mass PCR testing of their workforce was to be carried out.

Forty-two of 290 workers (14%) tested positive on initial mass testing, 25 (or 59%) of whom worked in the boning hall (in which there were a total of 64 workers deployed on shift at that time). Over a two month period, 111 asymptomatic workers, representing over one third of the workforce, tested PCR positive (Figure 2). They ranged in age from 22 to 64 years with a mean age of 40 years. Ninety-two of these workers when asked for details of possible transmission by the MPP management, were of the opinion that they had acquired infection outside of work. In total, 60 of the positive cases had been working in the boning hall, with a greater proportion of those cases occurring in workers on the night shift (19.8% of total shift workers), than on the day shift (9% of total shift workers).

### 3.2 Implementation of Risk Mitigation Measures

The initial COVID-19 mitigation measures implemented at this plant included checking of body temperature of all persons at point of entry, and other public health measures advised at that time, such as attention to hand hygiene and physical distancing. Public health advice evolved to include wearing PPE (firstly visors, then face masks) but not until mid-2020, i.e. several months after the outbreak had occurred in this plant. As concerns about the vulnerability of MPPs grew, the collective experience of ERTs within plants, including in the study plant, had a progressively greater influence on the specific adaptation of these general recommendations in specific plant settings. Further specific measures that were adapted to control COVID-19 included increased data collection, restrictions on visitors to the plant, definition of “pods” within the workforce, on-site presence of essential workers only, installation of Perspex dividers between some working stations, and increased vigilance on compliance with distancing on the floor and in rest areas, as well as mask wearing.

At the end of quarter 1 2020 and into quarter 2, guidelines were issued by the public health authorities for food business operators to aid in the management of the risk presented by COVID-19, *via* the employers’ representative body, Meat Industry Ireland (MII). These were augmented and adapted by plant management, who considered what could be done to mitigate risk within the specific environment of the MPP. These measures were progressively modified by the ERT in line with the growing knowledge of SARS-CoV-2, and the associated epidemiology of the disease. A summary of the main measures implemented, along with a timeline, is presented in Table 3.

During spring and early summer 2020 the MPP implemented a series of specific measures aimed at preventing workers from acquiring SARS-CoV-2 infection in the community (i.e. outside the workplace). These measures included communications, education, and travel protocols. Letters were issued to workers to inform them of the reasons for the plant being considered a high-risk setting for COVID-19, about the availability of an occupational health service, and social welfare provisions. Educational risk mitigation measures included posters (in several languages), explanation of COVID-19 symptoms, information about self-isolation and mitigation of risk in the community, handwashing signage (in several languages), and a COVID-19 induction programme. Travel risk mitigation included requesting staff not to share car journeys or travel with pod members, pod questionnaires and provision of PPE such as gloves, masks, and sanitizer for personal use when travelling to and from the plant.

During Spring 2020 the MPP also implemented some specific measures to prevent an infected individual from entering the plant. These included controls at point of entry, a return-to-work protocol, and restrictions on entry of non-staff members. Entry point controls included the use of hand sanitizer, temperature screening, a COVID-19 questionnaire, and a “restricted entry” list. All staff entry cards were disabled to facilitate the entry controls, and the restrictions were highlighted by prominent signage. On return to work after a period of absence, staff were given a questionnaire, which included questions about their experience of potential COVID-19 symptoms, as well as other symptoms. These Return to Work (RTW) protocols followed guidance issued by the public health authorities, and, where relevant, GP clearance for RTW. The following actions were introduced to restrict the entry of non-staff members; regulations preventing staff contact with farmers bringing animals to the plant, a prohibition on non-essential visitors and a COVID-19 questionnaire for essential visitors, such as maintenance contractors.

A wide variety of risk mitigation measures were directed at preventing the risk of within-plant transmission of infection. These included increased hygiene measures, limiting staff numbers at workstations and within common areas, provision of PPE, senior management response, positive cases response, and coordinated response from public health authorities in the form of local outbreak control teams. Specific measures to prevent infection within the factory including provision of hand sanitizer, staggered break times, and physical barriers between workstations in the boning hall. Additionally, the wearing of face masks was made mandatory in late Spring, and by the following month there was a reduction in the number of workers allowed into the canteen area. Additional protocols were put in place for potential COVID-19 related events including referrals for Covid-19 PCR testing, pods sent home to self-isolate, and 14-days isolation if testing positive.

### 3.3. Environmental (Air Quality) Assessment

The average CO_2_ concentrations and particle number concentrations measured during the daytime working shifts (between 07:00 and 15:00) in the boning hall over a three-day period and the abattoir over a two-day period are shown in Figure 3. The CO_2_ concentration in the boning hall showed a gradual increase throughout each working shift shift but dropped sharply on two occasions, corresponding to break times when the workers left the hall. However, when the workers returned, the amount of CO_2_ quickly increased again. The average count of fluorescent and total particles of less than 2.5 μm diameter (PM_2.5_) also gradually increased over the course of each working shift in the boning hall.

**Figure 3.**
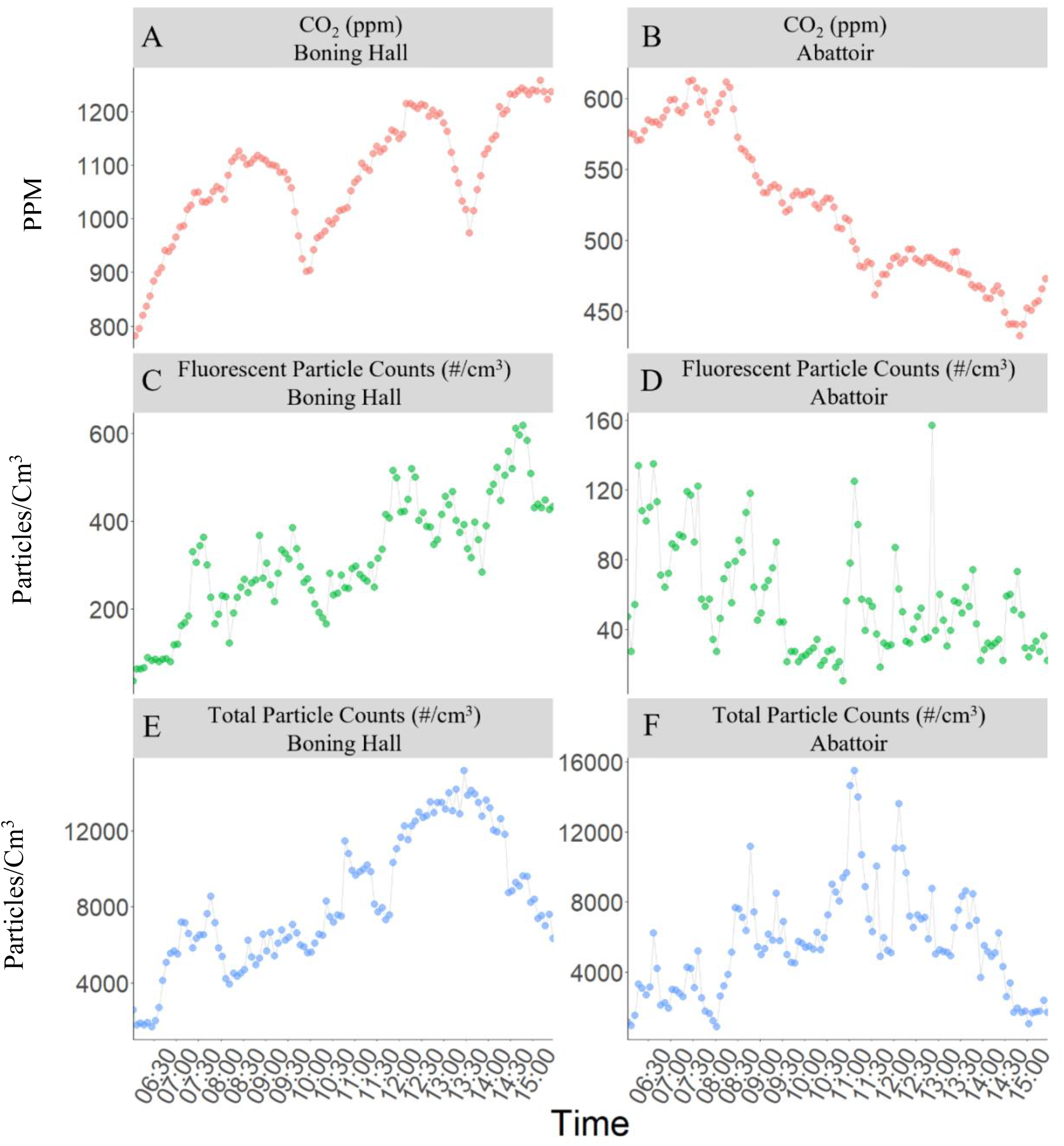
Average CO_2_ concentrations (A, B), Flourescent particle number concentration (C,D) and Total Particle Counts (E, F) measured during the daytime working shifts (07:00-15:00) in the boning hall (over 3 days) and abattoir (over 2 days).

In contrast, the CO_2_ concentration in the abattoir showed a marked decrease during the working shift and at one point reached levels close to typical outdoor values (ca. 415 ppm). The total particle concentration in the abattoir fluctuated greatly during the working day. However, the number of fluorescent particles was low and showed no significant change over time. The average air temperatures were 10 °C in the boning hall and 18 °C in the abattoir. The relative humidity was higher on average in the abattoir (71%) than in the boning hall (66%).

The air handling regimes differed between both areas of the plant - warm, humid air was continuously extracted from the abattoir whereas chilled air was being continuously recirculated within the boning hall. Based on the volumes of air extracted, it was estimated that the abattoir undergoes eight air changes per hour (ACH). In contrast, there is no extraction of air from the boning hall (the chilled air within being continuously circulated) and air exchange was limited to that which occurred through openings into adjacent areas of the plant. Based on the decay of CO_2_ concentration during the two break periods (Figure 3), it is estimated that the boning hall undergoes 0.4-0.5 ACH. This is indicative of poor ventilation and explains the gradual rise in both particle counts and CO_2_ over each working shift. Furthermore, the accumulation of fluorescent particles in this area suggests that bioaerosols are present in the air and remain airborne for the duration of the working shift.

### 3.4 Environmental Risk Mitigation Measures

The impact of installing air filtration units on average particle counts measured by the WIBS-4a during the different operational modes of the filtration unit is provided in Table 2. The results are based on measurements made during working shifts where the filtration unit was not operational (18 hours), operating at full capacity (5 hours) and 50% capacity (45 hours). The filtration unit reduced the total number of particles < 2.5 μm by 75% and 86% when operating at half and full capacity respectively. Similar results were obtained for the fluorescent fraction of particles, as well as particles > 2.5 μm.

**Table 2.** Average particle counts (#/cm^3^) measured by the WIBS during the different operational modes of the filtration unit in the boning hall and the associated reduction when compared to no filtration.

| Type | Size ($\mu m$) | Filtration Capacity | | | Percentage Change | |
| --- | --- | --- | --- | --- | --- | --- |
|  |  | 100% | 50% | None |  |  |
| | | Mean ± SD | | | 100% capacity | 50% capacity |
| WIBS_ | < 2.5 | 3508 ± 1744 | 6454 ± 4048 | 25522 ± 10804 | -86.30% | -75.00% |
| Total | > 2.5 | 569 ± 191 | 769 ± 369 | 2743 ± 791 | -79.30% | -72.00% |
| WIBS_ | < 2.5 | 241 ± 90 | 315 ± 172 | 1297 ± 338 | -81.40% | -75.70% |
| Fluorescent | > 2.5 | 235 ± 120 | 316 ± 149 | 834 ± 169 | -71.80% | -62.10% |

**Table 3.** Progressive timing of Implementation of Risk Mitigation Measures, classified as Prevention of Infection, Prevention of Entry of Infected Individuals, Prevention of Spread of Infection, and High-Level Management Response.

| Prevention of infection of staff with Covid |  |  |  |  |  |
| --- | --- | --- | --- | --- | --- |
|  | Quarter 1 2020 |  | Quarter 2 2020 |  |  |
| Communication | Letter to staff<br>High risk conditions and need to go to occupational health doctor |  |  | Information about social welfare compensation<br>Letter to staff | Whatsapp group – daily messaging<br>Factory wide communication evening with CC, gardai and managements |
| Education | Posters – several languages<br>Covid signs and Controls explained in letter<br>Staff on each line educated on Covid 19 signs – when not to come and contact GP |  |  | Information about self isolation and mitigation of risk in community<br>Covid Signs and controls explained in letter | Handwashing signage in different languages<br>Covid19 tailored induction introduced |
| Travel risk mitigation | Request to not carpool<br>Or travel with zone members | Pod questionnaire |  |  | Provided gloves, masks and sanitizer |

|  | Quarter 1 2020 |  | Quarter 2 2020 |  |
| --- | --- | --- | --- | --- |
|  | Early March | Late March | Early April | Late April |
| Entry point control | Hand sanitizer<br>Temp screening<br>Covid symptom survey | Disabled all staff entry cards<br>Restriction list, for RTW illness/travel<br>Covid signage put in place |  |  |
| Return to work protocol | RTW protocols introduced<br>RTW staff, separate flow chart questionnaire | Covid related symptoms – HSE rtw policy<br>Non-covid symptoms – GP clearance for RTW |  |  |
| Restrictions of non-factory staff | Staff contact with haulers/farmers stopped | VI restrictions<br>Refusal of visitors<br>Restriction on hauler/farmer movement on site<br>Covid survey for essential non-factory staff |  |  |

| Prevention of spread of infection within the factory |  |  |  |  |  |
| --- | --- | --- | --- | --- | --- |
|  | Quarter 1 2020 |  | Quarter 2 2020 |  |  |
| Increased Hygiene | Hand sanitizer factory wide<br>Increased cleaning of toilets, locker rooms<br>Increased cleaning of corridors and high touch points<br>Disinfection of canteen between breaks<br>Cleaning of office high touch points<br>Common area Covid cleaning SOP Issue 1 | Induction room to be sanitized before and after use<br>Fogging weekly of factory floor lines |  |  | Covid Cleaning SOP Issue 6 – far more detailed and area specific, with training for cleaners included. |
| Managing staff numbers | Staggered break times to prevent congregation<br>Zoning on lines<br>Office staff who can work from home sent home<br>Extended break between boning hall shifts | Pod questionnaire – area on line, travel and living arrangements.<br>Specific restriction on managers, supervisors and charge-hands | Addition of Chill 4 to both boning shifts – more distance<br>Movement of staff between lines must remain in Pod |  |  |
| Managing staff work place | Reposition of staff 2m distance boning hall<br>Pod allocation for areas where 2m distance was not possible<br>Other lines – 2m distance normally in place<br>Relocation of office staff to allow distancing |  | Blue boards, physical barrier in boning hall<br>Slowed lamb line to allow social distance |  | Curtain – physical barrier in boning hall |
| Managing common areas | Designated canteen areas according to zone<br>Designated smoking areas according to zone<br>Policing of canteen and locker areas<br>Staggered break times for toilets | Designated times for dining for all staff<br>Advice signage about covid smoking etiquette | Floor markings<br>Physical barrier for canteen staff<br>Covid supervisors put in place – social distancing and seating | Cooking in canteen ceased<br>Queueing system modified | Reduced numbers in canteen further (80-30)<br>Marquee with social distancing for overflow |
| PPE |  | Masks made available to staff |  | Mandatory mask wearing |  |

| Prevention spread of infection with the factory MANAGEMENT RESPONSE |  |  |  |  |
| --- | --- | --- | --- | --- |
|  | Quarter 1 2020 |  | Quarter 2 2020 |  |
| High level response | Establishment of Covid Committee.<br>SOP for site management developed | Assignment of responsibility to designated aspects of Covid control |  |  |
| Positive cases response |  | Formation of protocol for Covid possible events:<br>o Symptomatic at work<br>o Symptomatic at home<br>o Close contact<br>Note – assumption transmission needed case to be symptomatic |  | Addition of protocol for Covid possible events:<br>o Referred for Covid test<br>o Referred but not done<br>o Covid positive<br>Rapid identification of Pod and Zone<br>Pod sent home for isolation<br>14 days isolation if + |
| Coordinated LOCT response |  |  |  | HSE suggest mass testing |

### 3.5 Visual assessment and physical measurement of operational risk factors

In the boning hall, workers were typically in stationary positions and operated less than 1 m from each other but were separated by panels and were provided with surgical masks and visors. Social interaction was limited due to the physical barriers. Human occupancy was much higher in the boning hall than in the abattoir – with three times less floor area per person (5.1 m^3^ compared to 15.2 m^3^) and four times less airspace per person (25.5 m^3^ compared to 108.2 m^3^).

### 3.6 Post-outbreak events

From mid-2020, and the following 12 months, the factory had no COVID-19 outbreaks, and no linked cases dating from 198 days from the last detected case. This time represents 39,600 person days free of any new within-plant transmitted cases. In addition, between 0 and 3 workers tested PCR positive for SARS-CoV-2 at each serial test of the workforce, performed at four-weekly intervals from Quarter 2 2020 to Quarter 2 2021. However, none of the positive cases were linked and none were considered to constitute an outbreak. The risk mitigation measures, as outlined, have been consistently maintained.

## 4.0 Discussion

COVID-19 transmission within MPPs has occurred across the globe, especially in countries where the meat industry is consolidated, and large processing facilities are operated (1,2,3,12). Temperature, humidity, and air circulation are all known to play a significant role in the transmission and stability of SARS-CoV-2 (5,7,8,9). Susceptibility of MPPs to COVID-19 outbreaks has been attributed to many different factors both outside and within the plants (4, 15). We present here a retrospective study of an outbreak in a MPP in Ireland documenting and evaluating risk factors within the plant and a series of risk mitigation measures used by plant management. Our findings add to the existing literature on MPP Covid-19 outbreaks in Germany, USA, Italy, and Ireland.

The risk mitigation measures implemented at this plant included controls at the point of entry (questionnaire and temperature checks), widespread signage highlighting COVID-19 risk mitigation measures (in multiple languages), a greater emphasis on hand hygiene including the increased use of sanitizer, installation of physical barriers between adjacent workers, compulsory use of face masks and restrictions on occupancy levels (including staggered work shifts and work breaks).

These measures were based on generic public health recommendations, but were refined incrementally after the outbreak had occurred, based on the first-hand knowledge of operational risk factors, and increasing specific expertise in COVID 19 mitigation in a meat plant setting, through the plant’s ERT. In the early days of the pandemic, due to the weak evidence base and need for quick decision making, public health advice sometimes lagged behind the measures employed by the MPP, with official recommendations for MPPs only issued from May onwards, and with limited updating. For example, initial advice cited in the first risk assessment declared asymptomatic people not to be contagious. Initial advice favoured plastic visors and discouraged the wearing of face masks, and public health advice influenced the installation of physical dividers in the boning hall, without evidence of efficacy in preventing virus transmission. In addition, the practical difficulties in maintaining a physical distance of 2 m in an MPP operational setting, with significant ambient noise, could not be fully appreciated by those unfamiliar with these environments, as was clearly documented in our study.

These public health measures were primarily aimed at mitigating droplet or contact transmission of SARS-CoV-2. In contrast, there was relatively little attention paid to the specific risks of aerosol transmission, which occurs by inhalation of suspended respiratory particles that contain infectious virus, both at short-range and long-range (> 2 m from the source). During 2020, reports were emerging of long-range aerosol transmission of SARS-CoV-2, particularly in crowded and poorly ventilated indoor settings, which are now widely accepted (15). Gunther et al (12) clearly demonstrated the potential for aerosol transmission of SARS-CoV-2 over distances as much as 8 metres in a MPP, and also highlighted the increased risk in meat cutting rooms which, for food safety reasons, have to be maintained at low temperature, and this is typically achieved by recirculation of chilled air.

The operational conditions in the boning hall of the MPP investigated during this study is very similar to that reported by Gunther et al (12) and this is the area of the plant where the largest number of PCR-positive cases were detected among workers. Two risk factors combine within boning halls to provide an environment that is highly favourable for aerosol transmission of SARS-CoV-2 as well as other airborne infections – these being high occupancy and poor ventilation. Boning halls tend to be more densely populated than other work areas – that in the present study had three to four times the occupancy of the abattoir in the same plant. Meanwhile air quality results in this study, showing a gradual increase in CO_2_ levels and in suspended bio-aerosols over the course of a working shift within the boning hall (but not in the abattoir) confirm that this area of the plant is poorly ventilated. Our results indicate that CO_2_ concentration is closely corelated with occupancy levels. Bioaerosols could have originated from exhaled respiratory particles emitted by the workers (which could contain the SARS-CoV-2 virus) or from tiny particles of meat and bone that became airborne.

The ventilation regime currently implemented in boning halls is deliberate. EU food hygiene legislation requires that meat cutting rooms are maintained at a temperature of less than 12 °C (to ensure that red meat is kept at below 7 °C). Up until now, the international industry norm to achieve these working conditions in meat cutting rooms has been to continuously recirculate chilled air with little or no fresh air intake or filtration.^3^ However, this legislative requirement introduced in the wake of the BSE crisis, predates some fundamental changes in the efficiency of meat processing. Typically, meat cutting is now a much more streamlined, assembly-line process with greater throughput and shorter chill-to-chill transit times than heretofore. EU legislation already allows for alternatives (to a working temperature of less than 12 degrees) to achieve the same effect (of keeping red meat at less than 7 degrees). This legislative proviso coupled with current more efficient processes provide scope to explore if meat cutting could be performed in rooms operated at a higher ambient temperature without compromising on food safety. If such an alternative were validated, there would be less of a requirement to recirculate air and more fresh air could be introduced to meat cutting rooms without significantly increasing the energy costs or carbon footprint of meat processing. Work to investigate if modifying this requirement, while still providing for food safety considerations, is underway.

Carbon dioxide concentrations are routinely used as an indicator for the adequacy of ventilation in buildings and low-cost sensors can allow for quick and easy measurement of CO_2_. While there is currently no legal limit prescribed for CO_2_ concentration in indoor settings in Ireland and there are very few examples of such limits imposed in other jurisdictions, Belgium has recently introduced a legal limit of 900 ppm, with levels above 1200 ppm deemed a safety breach. Ireland has occupant building ventilation regulations requiring the minimum capacity of a centralized continuous mechanical extract ventilation system to give 10 L of air per second per person^4^.

In indoor settings where air exchange is not sufficient, the physical removal of aerosol particles from air by filtration, provides an alternative means of improving indoor air quality. The CDC ^5^ and WHO^6^ recognise portable, industrial-grade, HEPA (High Efficiency Particle Air) filtration devices as a supplemental means of increasing the effective number of air changes per hour (ACH) in controlled environments. Furthermore, HEPA filtration devices have been successfully used for decades to reduce the concentrations of airborne particles, including the removal of bacteria and viruses from indoor air (16). The installation and operation of a large HEPA filtration unit within the boning hall investigated in this study was shown to reduce the build-up of airborne particles over the course of a working shift. Significant reductions in particle counts were achieved even when the filtration unit was only operating at half its functional capacity. While a further reduction could be achieved if the unit was operated at full capacity, this was associated with a “wind chill” effect and was excessively noisy, negatively impacting on worker comfort and therefore unlikely to be acceptable on a long-term basis. Further work to assess the importance of such measures will be important in mitigating ongoing risk.

### 4.1 Conclusions

This study provides a significant insight into specific operational and environmental factors pertaining to COVID-19 spread within MPPs. It is clear that working and environmental conditions are conducive to SARS-CoV-2 transmission, and that strict risk mitigation measures are required to protect workers, and, by extension, the local community. Specific attention should be directed towards risk mitigation (ventilation, air extraction and filtration) in boning halls and other environments where bioaerosol build-up is likely. Our findings emphasise the important role of plant management and the expertise of those working in the sector to translate and apply general public health recommendations appropriately to the specific environments in MPP, and to customize risk mitigation measures and coordinate their implementation so that they are maximally effective. This sector is accustomed to maintaining controls for food safety requirements, and thus is ideally positioned to use this expertise in protection of occupational and public health. The benefits of co-operation between public health authorities, experts within the industry, and those with expertise in veterinary public health were highlighted in this study. In addition, the findings of our study confirm the important role of ventilation and air quality measurements in reducing the threat posed by airborne transmission of SARS-CoV-2 in workplace settings with high occupancy levels. This study emphasises, above all, the value of involving the general public, businesses, and communities, as partners, rather than passive subjects, in the public health response. The management response to the outbreak documented here is an exemplar of successful implementation of this approach.

There are limitations to our study which it is important to acknowledge. We have presented a retrospective analysis carried out in one meat processing plant, involving an investigation of events that occurred months previously. It is important to note that the data collected was provided by the local management team in the meat plant some months after the events described and was based on details that workers disclosed to their employer at the time of the outbreak and this may have introduced different types of bias, including recall bias. The bioaerosol measurements reported here were also limited to a total of five days in two different areas of the plant.

Further studies of COVID-19 in MPPs are indicated as a public good - such work is progressing in Ireland, supported by the State, including the public health authorities, by the meat industry and by all other relevant stakeholders.

## Data Availability

Raw data files are maintained in encrypted form by the project consortium and are available on request from the authors

## Author Contributions

Data Collection – NW, MF, SH, JW, VJ, GM, VC, DS

Data Analysis – NW, MF, SH, JW, MP, VJ, VD, CP, GM, DS

Manuscript Drafting – All authors Manuscript

Editing – All authors

## Acknowledgements

We are grateful to Meat Industry Ireland and their member companies for supporting this work and for ongoing studies under the Science Foundation Ireland grant listed above. We also thank colleagues from the Health and Safety Authority, and in particular Jason Murray, for their advice and assistance.

## Footnotes

1 https://www.theguardian.com/australia-news/2020/jul/22/coronavirus-clusters-why-meatworks-are-at-the-frontline-of-australias-second-wave

2 https://www.bordbia.ie/industry/irish-sector-profiles/

3 [Regulation (EC) No 853/2004](https://www.fsai.ie/uploadedFiles/Consol_Reg853_2004(1).pdf)

4 Department of Housing, 2021

5 https://www.cdc.gov/coronavirus/2019-ncov/community/ventilation.html#refphf.

6 https://apps.who.int/iris/bitstream/handle/10665/339857/9789240021280-eng.pdf?sequence=1&isAllowed=y

